# Emergence of the Delta Variant and risk of SARS-CoV-2 infection in secondary school students and staff: prospective surveillance in 18 schools, England

**DOI:** 10.1101/2021.12.10.21267583

**Authors:** Shamez N. Ladhani, Georgina Ireland, Frances Baawuah, Joanne Beckmann, Ifeanyichukwu O Okike, Shazaad Ahmad, Joanna Garstang, Andrew J Brent, Bernadette Brent, Felicity Aiano, Zahin Amin-Chowdhury, Meaghan Kall, Ray Borrow, Ezra Linley, Maria Zambon, John Poh, Lenesha Warrener, Angie Lackenby, Joanna Ellis, Gayatri Amirthalingam, Kevin E Brown, Mary E Ramsay

## Abstract

**Background:** The role of educational settings on SARS-CoV-2 infection and transmission remains controversial. We investigated SARS-CoV-2 infection, seroprevalence and seroconversions rates in secondary schools during the 2020/21 academic year, which included the emergence of the more transmissible Alpha and Delta variants, in England.

**Methods:** The UK Health Security Agency (UKHSA) initiated prospective surveillance in 18 urban English secondary schools. Participants had nasal swabs for SARS-CoV-2 RT-PCR and blood sampling for SARS-CoV-2 Nucleoprotein and Spike protein antibodies at the start (Round 1: September-October 2020) and end (Round 2: December 2021) of the autumn term, when schools reopened after national lockdown was imposed in January 2021 (Round 3: March-April) and end of the academic year (Round 4: May-July).

**Findings:** We enrolled 2,314 participants (1277 students, 1037 staff). In-school testing identified 31 PCR-positive participants (20 students, 11 staff). Another 247 confirmed cases (112 students, 135 staff) were identified after linkage with national surveillance data, giving an overall positivity rate of 12.0% (278/2313; staff [14.1%, 146/1037] vs students [10.3%, 132/1276; p=0.006). Nucleoprotein-antibody seroprevalence increased for students and staff between Rounds 1-3 but changed little in Round 4, when the Delta variant was the dominant circulating strain. Overall, Nucleoprotein-antibody seroconversion was 18.4% (137/744) in staff and 18.8% (146/778) in students, while Spike-antibody seroconversion was higher in staff (72.8% (525/721) than students (21.3%, 163/764) because of vaccination.

**Interpretation:** SARS-CoV-2 infection and transmission in secondary schools remained low when community infection rates were low because of national lockdown, even after the emergence of the Delta variant

**Funding:** DHSC

## Introduction

Children have a lower risk of severe or fatal COVID-19 compared to adults.^1^ Early in the pandemic, however, schools were closed as part of national lockdown in most parts of the world, including the United Kingdom, due to uncertainties of their role in transmission. As COVID-19 cases declined, some UK primary and secondary school years were partially reopened during June-July 2020 with strict infection control measures, physical distancing and smaller class sizes clustered into distinct bubbles that did not interact with other bubbles.^2^ SARS-CoV-2 infections remained low, with few outbreaks reported in educational settings, leading to the full reopening of in-person teaching in schools across the UK from September 2021.^2,3^

In England, community cases started increasing across all age-groups from August 2020 and continued to increase at the start of the first academic term of 2020/21 (Sep-Dec), with cases in children lagging behind adults until another national lockdown was imposed on 05 November 2020, although schools remained open during this period.^4^ The emergence and rapid spread of the Alpha variant in the UK from mid-November 2020 led to a third national lockdown in January 2021, including school closures. Cases declined rapidly across all age-groups during January and February 2021 and remained low until mid-May 2021.^5^

From 08 March 2021, schools were reopened for three weeks whilst the rest of the country remained in lockdown and then closed for the Easter holidays. Summer term began in mid-April to mid-July 2021, with a one week half-term break in May. During this time, England gradually eased out of lockdown. While cases due to the Alpha variant continued to decline rapidly from January 2021, the Delta (B.1.617.2) variant emerged in the UK in mid-March 2021 and became the most prevalent variant in England from mid-May 2021.^6,7^

Nationally, community SARS-CoV-2 infection remained low until 17 May 2021 (week 20) and then started increasing rapidly, mainly in older teenagers and younger adults, who were unvaccinated at the time.^5,8^ Older adults were protected through COVID-19 vaccination, which began in December 2020, and by mid-June 2021, the vaccine was being offered all adults from 18 years of age. ^9^ The combination of national lockdown, testing patterns (which included twice-weekly home testing with lateral flow device (LFD) tests for secondary school students) and COVID-19 vaccination for adults changed the epidemiology of SARS-CoV-2 infection in England, such that 10-19 year-olds had the highest rates of infection after schools reopened on 08 March 2021.^10^

In England, The UK Health Security Agency (UKHSA) (formally known as Public Health England) has been conducting enhanced surveillance of COVID-19 in selected secondary schools across England since September 2021. This included blood sampling for SARS-CoV-2 antibodies, which captures both symptomatic and asymptomatic SARS-CoV-2 infection, in staff and students.^3^ Three earlier rounds of testing found that, by March 2021, 36% of students and 32% of staff had nucleocapsid protein (N) antibodies indicating prior infection, with an additional third of staff members having spike protein (S) antibodies through vaccination.^3,11^ Between May and July 2021, we completed a final round of testing in secondary schools when the Delta variant was prevalent across England and community cases were increasing nationally.^5^ Here we report the SARS-CoV-2 infection and antibody seroprevalence in secondary schools between May and July 2021 and assessed trends throughout the academic year compared with national infection surveillance and testing data in England.

## Methods

The COVID-19 Surveillance in Secondary School KIDs (sKIDsPLUS) protocol is available online (https://www.gov.uk/guidance/covid-19-paediatric-surveillance),^12^ and results for the first three rounds of testing have been published.^3,11^ The study involved testing secondary school students for SARS-CoV-2 infection and antibodies at the start (Round 1: 22 September-17 October 2020) and end (Round 2: 3-17 December 2020) of the autumn term of the 2020/21 academic year, when the schools reopened in March (Round 3: 23 March-21 April 2021) and at the end of the academic year (Round 4: 20 May-14 July 2021). Participants who were isolating during Round 4 testing were sent a Tasso+ blood self-sampling device at home, which was posted back to UKHSA for antibody testing.^13^ Secondary schools were approached in areas where a paediatric investigation team could be assembled: Derbyshire, West London, East London, Greater Manchester, Hertfordshire and Birmingham. Headteachers in participating schools emailed the study information pack to staff, parents of students aged <16 years and to students aged ≥16 years. Participants or their parent/guardian provided informed consent online via SnapSurvey, and completed a short questionnaire on COVID-19 symptoms and confirmed infection prior to the sampling day or shortly afterwards. Enrolment was open for new participants between Rounds 1 and 2, although 94 participants didn’t participate until Round 3. A team of clinicians, nurses, phlebotomists and administrative staff attended the school on sampling days and a nasal swab and blood sample was taken for each participant. Local anaesthetic cream was offered to all students before blood sampling.

### Laboratory testing

The swabs were tested by a triplex reverse transcription PCR (RT-PCR) assay for the detection of ORF1ab and E gene regions of SARS-CoV-2 with simultaneous detection of an exogenous internal control using the Applied Biosystems Quantstudio 7-flex thermocycler (ThermoFisher Scientific, UK).

The ORF1ab gene primers/probes published by the Chinese Centre for Disease Control and Prevention were combined with the E gene primers/probe published Corman *et al*., 2020.^14,15^ A positive RT-PCR result was reported to the participant, local investigator, head teacher and local UKHSA health protection team (HPT), typically within 48 hours of the sample being taken. The participant and household members self-isolated as per national guidance. Public health risk assessment was undertaken with the school to decide additional measures, including tracing.

In Rounds 1-3, serology was performed on the Abbott Architect, using a chemiluminescent microparticle immunoglobulin G (IgG) immunoassay targeting the nucleoprotein (N) (SARS-CoV-2 IgG, Abbott Commerce Chicago, USA) with a seropositivity cut-off value of 0.8 (henceforth referred to as Abbott N assay).^16^ This assay was shown to detect SARS-CoV-2 N-antibodies as early as 7 days post-symptom onset and is, therefore, particularly useful for rapidly assessing SARS-CoV-2 antibody seroconversion (negative to positive antibodies) between testing rounds.^17^ Detectable N-antibody with the Abbott N-assay, however, falls rapidly after >3 months following SARS-CoV-2 infection, more than other commercial and in-house N-antibody assays, indicating that this is a characteristic of the assay rather than actual loss of N antibodies in most participants.^18^ Where sufficient serum was available, samples from all rounds were additionally tested for nucleoprotein and spike (S) protein antibodies on the Roche Elecsys Anti-SARS-CoV-2 N assay and Elecsys Anti-SARS-CoV-2 S assay (henceforth referred to as Roche N and Roche S assays). In Round 4, serology was performed on Roche N and S assays, due to their higher sensitivity and specificity and apparent antibody waning with the Abbott N assay, and where sufficient sample remained, samples were also tested on the Abbott N assay.^18^

### Statistical Analysis

Participants were linked to national laboratory report of SARS-CoV-2 PCR and LFD tests (SGSS) to identify diagnoses between testing rounds. A combination of unique personal NHS number (obtained by linking with the Personal Demographic Service, an online national electronic database containing demographic information for all NHS-registered individuals), full name, sex, date of birth and postcode of residence was used.^19^

Data were managed in R-Studio and Microsoft Access and analysed in Stata SE (version 15.1). Participants were classified as included in each round if they provided a blood sample or swab in that round. Data that did not follow a normal distribution are described as median with interquartile ranges. Categorical data are described as proportions and compared with the Chi^2^ test.

Results from the Roche N and S assay were used to present seroprevalence and estimate seroconversion rates. However, in Rounds 1 and 2, 9.7% (170/1754) and 5.9% (105/1766) persons, respectively, had insufficient remaining serum for testing on the Roche N and Roche S assays, the majority of whom were Abbott N positive. For these people, Abbott N results were used to supplement Roche N and S results. SARS-CoV-2 infection rate and antibody seroprevalence, with 95% confidence intervals (CI), were compared between secondary school students and staff. For comparison with community seroprevalence, three-week average seroprevalence data from UKHSA and NHS Blood and Transplant (NHS BT) serosurveillance of blood donors were obtained for each school area in sKIDsPLUS. Donor sera are tested for N and S-antibodies on the Roche assays and, for this analysis, student and staff data was compared to seroprevalence in 18-30 year olds and 18-64 year old donors, respectively. In Round 4, student Roche S seropositivity do not have a comparator because vaccination rates were high in the comparator adult age-groups. Non-overlapping 95% CIs were used to assess statistical significance between student or staff rates and regional estimates.

Antibody seroconversion rates, using the Roche N and S antibody results and with 95% confidence intervals, were calculated for participants who were tested in two sequential rounds and were negative in their first round of testing.

### Ethics statement

The protocol was approved by PHE Research Ethics and Governance Group (reference NR0228; 24 August 2020).

## Results

During the 2020/21 academic year (September 2020 to July 2021), 2,314 participants (1,277 students: 1,037 staff) were enrolled and sampled at least once (**Table 1**). Overall 29.8% of students participated in all 4 rounds of testing, 38.4% in 3 rounds, 23.4% in 2 rounds and 8.4% in 1 round. For staff, the corresponding figures were 46.1%, 28.3%, 16.0% and 9.6%. Following Round 4, 102 students and 40 staff returned a home testing sample. Age was significant predictors of the number of rounds students and staff participated in (p=0.0015 and 0.0022 respectively), as well as school area (both p<0.0001) (**Supplementary table 1**). Participation was higher in Rounds 1 (n=1,825), 2 (n=1,842) and 3 (n=1,895) than in Round 4 (n=1,359) (**Table 2**). Median time between testing rounds was 9.4 (IQR:9-11) weeks between Rounds 1-2, 15.9 (IQR: 15.1-15.9) weeks between Rounds 2-3 and 14.1 (IQR: 14-15) weeks between Rounds 3-4.

**Table 1:**
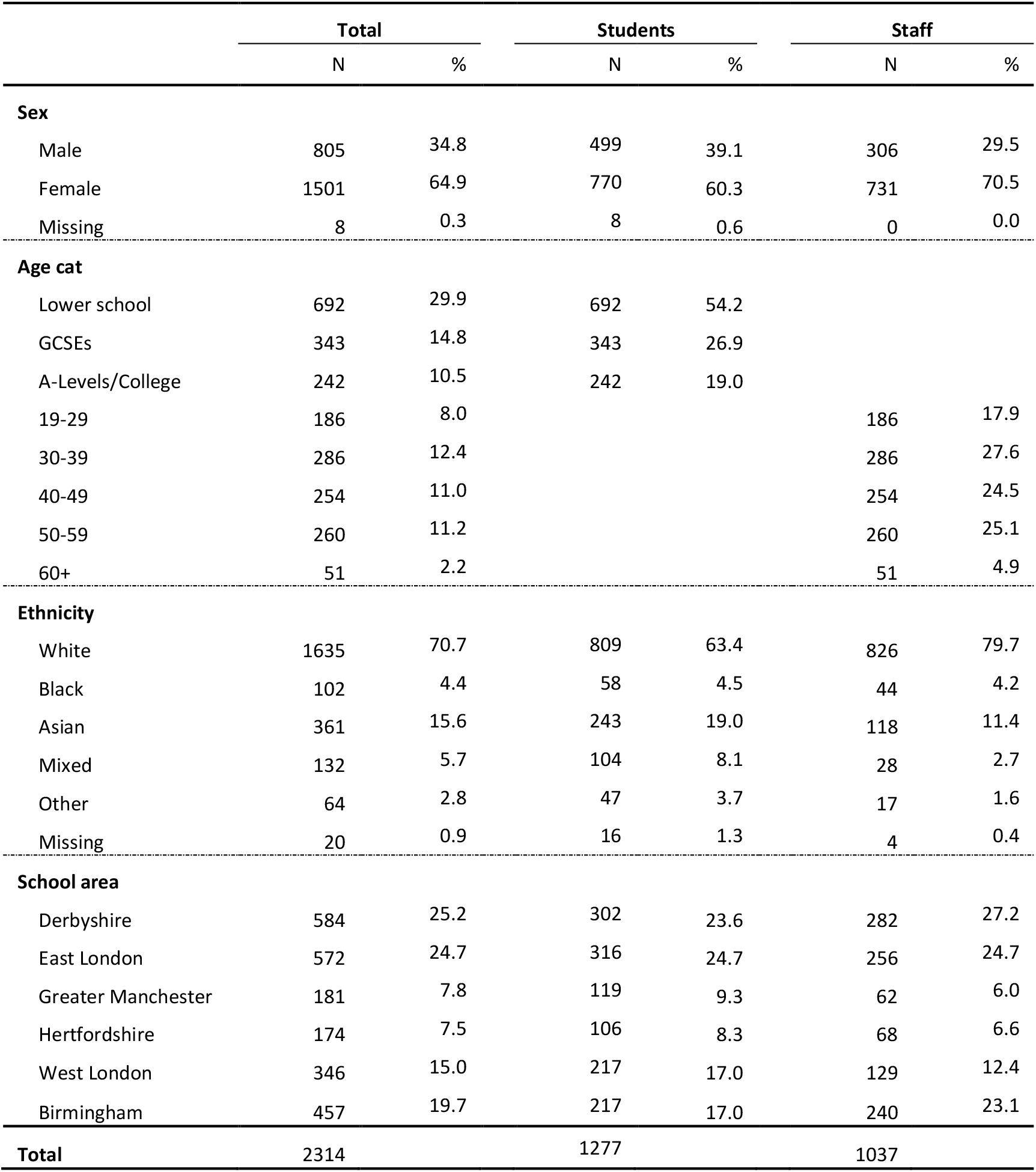
Characteristics of students and staff who participated in at least 1 round of sKIDs Plus.

**Table 2:** SARS-CoV-2 PCR and antibody (Roche N and Roche S) results over 4 testing rounds in sKIDs Plus participants.

|  | Round 1 |  |  | Round 2 |  |  | Round 3 |  |  | Round 4 |  |  | Tasso home sampling |  |  |
| --- | --- | --- | --- | --- | --- | --- | --- | --- | --- | --- | --- | --- | --- | --- | --- |
|  | Total | Students | Staff | Total | Students | Staff | Total | Students | Staff | Total | Students | Staff | Total | Students | Staff |
| *PCR results* |  |  |  |  |  |  |  |  |  |  |  |  |  |  |  |
| Negative | 1,817<br>(99.6) | 943<br>(99.5) | 874<br>(99.8) | 1,817<br>(99.1) | 938<br>(98.9) | 879<br>(99.2) | 1,884<br>(99.9) | 1,092<br>(99.8) | 792<br>(100.0) | 1,339<br>(99.6) | 689<br>(99.6) | 650<br>(99.7) |  |  |  |
| Positive | 7 (0.4) | 5 (0.5) | 2 (0.2) | 17 (0.9) | 10 (1.1) | 7 (0.8) | 2 (0.1) | 2 (0.2) | 0 (0.0) | 5 (0.4) | 3 (0.4) | 2 (0.3) |  |  |  |
| PCR total | 1,824 | 948 | 876 | 1,834 | 948 | 886 | 1,886 | 1,094 | 792 | 1,344 | 692 | 652 |  |  |  |
| *Roche N: Antibody results* |  |  |  |  |  |  |  |  |  |  |  |  |  |  |  |
| Negative | 1467<br>(83.6) | 742<br>(83.0) | 725<br>(84.2) | 1387<br>(78.5) | 695<br>(77.8) | 692<br>(79.3) | 1185<br>(65.7) | 657<br>(63.8) | 528<br>(68.3) | 846<br>(65.4) | 404<br>(61.7) | 442<br>(69.2) | 86<br>(60.6) | 59<br>(57.8) | 27<br>(67.5) |
| Positive | 288<br>(16.4) | 152<br>(17.0) | 136<br>(15.8) | 379<br>(21.5) | 198<br>(22.2) | 181<br>(20.7) | 618<br>(34.3) | 373<br>(36.2) | 245<br>(31.7) | 448<br>(34.6) | 251<br>(38.3) | 197<br>(30.8) | 56<br>(39.4) | 43<br>(42.2) | 13<br>(32.5) |
| Roche N total | 1,755 | 894 | 861 | 1,766 | 893 | 873 | 1,803 | 1,030 | 773 | 1,294 | 655 | 639 | 142 | 102 | 40 |
| *Roche S: Antibody results* |  |  |  |  |  |  |  |  |  |  |  |  |  |  |  |
| Negative | 1459<br>(83.1) | 737<br>(82.4) | 722<br>(83.9) | 1369<br>(77.5) | 687<br>(76.9) | 682<br>(78.1) | 936<br>(51.9) | 625<br>(60.7) | 311<br>(40.2) | 398<br>(30.8) | 384<br>(58.6) | 14<br>(2.2) | 48<br>(33.8) | 46<br>(45.1) | * |
| Positive | 296<br>(16.9) | 157<br>(17.6) | 139<br>(16.1) | 397<br>(22.5) | 206<br>(23.1) | 191<br>(21.9) | 867<br>(48.1) | 405<br>(39.3) | 462<br>(59.8) | 896<br>(69.2) | 271<br>(41.4) | 625<br>(97.8) | 94<br>(66.2) | 56<br>(54.9) | 38<br>(95.0) |
| Roche S total | 1,755 | 894 | 861 | 1,766 | 893 | 873 | 1,803 | 1,030 | 773 | 1,294 | 655 | 639 | 142 | 102 | 40 |
| Round total | 1,825 | 948 | 877 | 1,842 | 950 | 892 | 1,895 | 1,100 | 795 | 1,359 | 700 | 659 | 142 | 102 | 40 |

Of Round 4 participants, 52.6% (368/700) of students and 58.9% (388/659) of staff completed the sampling questionnaire. Since schools reopened in March 2021, most students (95.0%; 344/362) and staff (84.3%; 327/388) reported attending school every day and most students (63.7%; 230/361) and staff (76.0%; 295/388) had not missed any days off school.

### RT-PCR Results

Over 4 rounds of nasal swab testing, 31 (1.3%; N=2,313) participants tested PCR-positive (20 students, 11 staff). Infection rates among participants varied over time and closely followed community infection rates (**Table 2, Supplementary Figure 2**). When linked to national testing data, an additional 247 (10.7%) participants (112 students, 135 staff) had SARS-CoV-2 diagnosed over the study period, equating to a combined total of 12.0% (278/2,313) of participants testing positive. Positivity was higher in staff than students (14.1%, 146/1,037; vs 10.3%, 132/1,276; p-value=0.006), and most cases were between September-December 2020 (61.9%), compared to 22.7% between January and schools reopening on 08 March 2021, and 15.1% from 08 March until the final testing round (max: 14 July 2021) (**Supplementary Figure 3a**).

Amongst participants who were not tested in Round 4, 2.5% (17/670) of students and 1.4% (6/424) of staff had confirmed SARS-CoV-2 infection reported to national surveillance in the period between 10 days prior and 6 days after the scheduled school test date. When combined with the results of those who participated in Round 4, this would equate to an overall positivity of 1.5% (20/1362) in students and 0.7% (8/1076) in staff at the final round of testing.

### Antibody prevalence

Roche N antibody seroprevalence, indicating prior infection, increased for students and staff between Rounds 1, 2 and 3 but changed little in Round 4 (**Figure 1, Table 2**). Roche S antibody prevalence, which measures both prior infection and vaccine-induced antibodies, was similar to N antibody seroprevalence in Rounds 1 and 2 for staff and in all rounds for students, but was higher for staff for Rounds 3 and 4 because of COVID-19 vaccination, reaching >95% for staff in all school regions by Round 4. N and S antibody seroprevalence was higher, although non-significantly, in the 142 participants home-tested for SARS-CoV-2 antibodies because they missed testing in school (**Table 2**). In Rounds 1-4 some differences in antibody prevalence between the school areas were observed, but rates were generally similar to contemporaneous community seroprevalence estimates (**Supplementary Table 4**).

**Figure 1:**
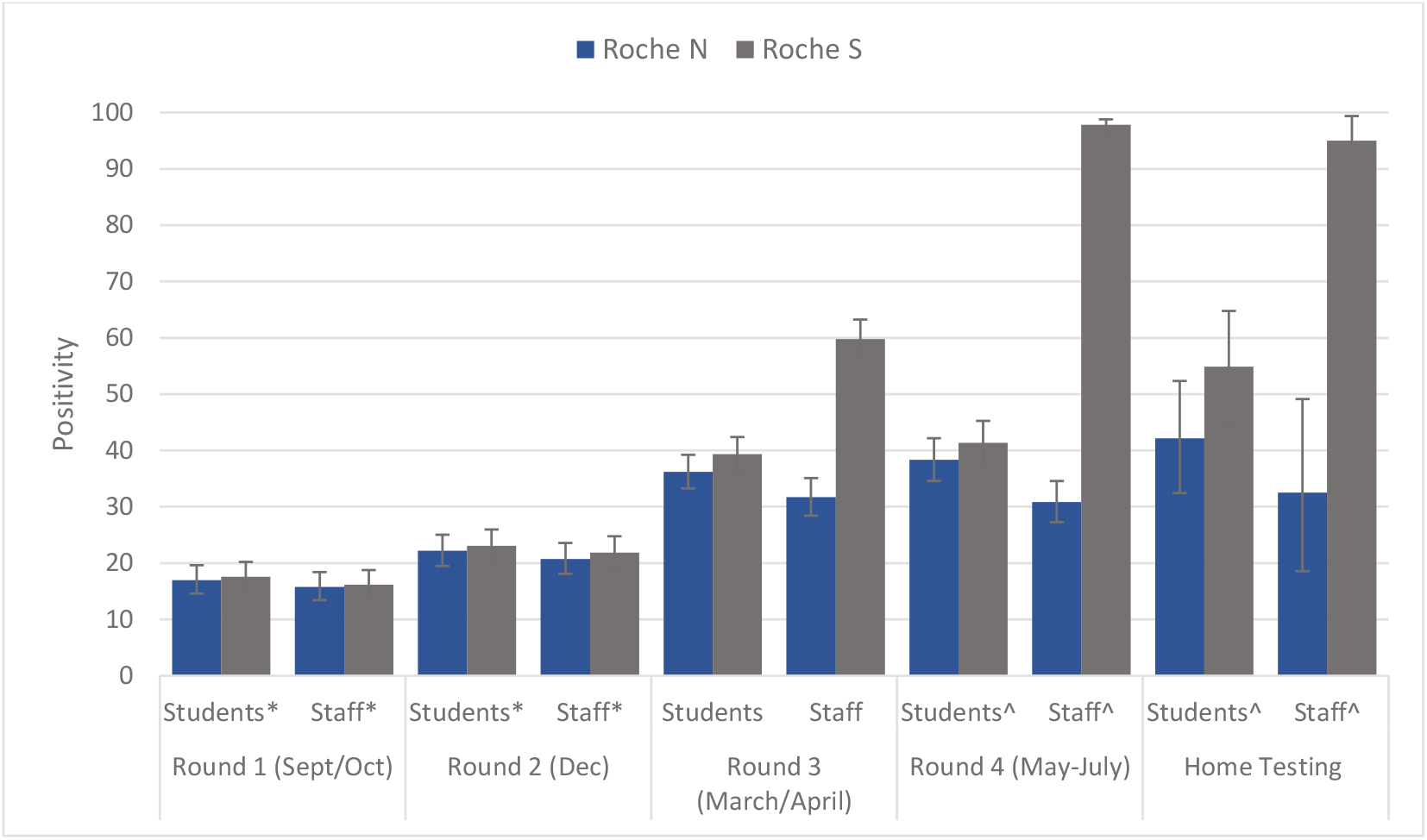
Roche N and S positivity, and 95% confidence intervals, in student and staff sKIDs PLUS participants in round 1, 2, 3 and 4 and via home sampling.

### Antibody seroconversion rates

Over Rounds 1-4, 18.8% (146/778) of students seroconverted based on their N antibody results and 21.3% (163/764) based on S antibody results, compared to 18.4% (137/744) and 72.8% (525/721) of staff. N antibody seroconversion rates were lowest between Rounds 3 and 4, at 1.44 (95% CI: 0.58-2.97) students per 1,000 weeks and 1.92 (95% CI: 0.92-3.53) staff per 1,000 weeks (**Figure 2 and Supplementary table 5**). Amongst N-antibody seroconverters who completed the questionnaire for Round 4 (52.9%; 9/17), six of nine individuals (66.7%) reported feeling unwell between rounds 3 and 4.

**Figure 2:**
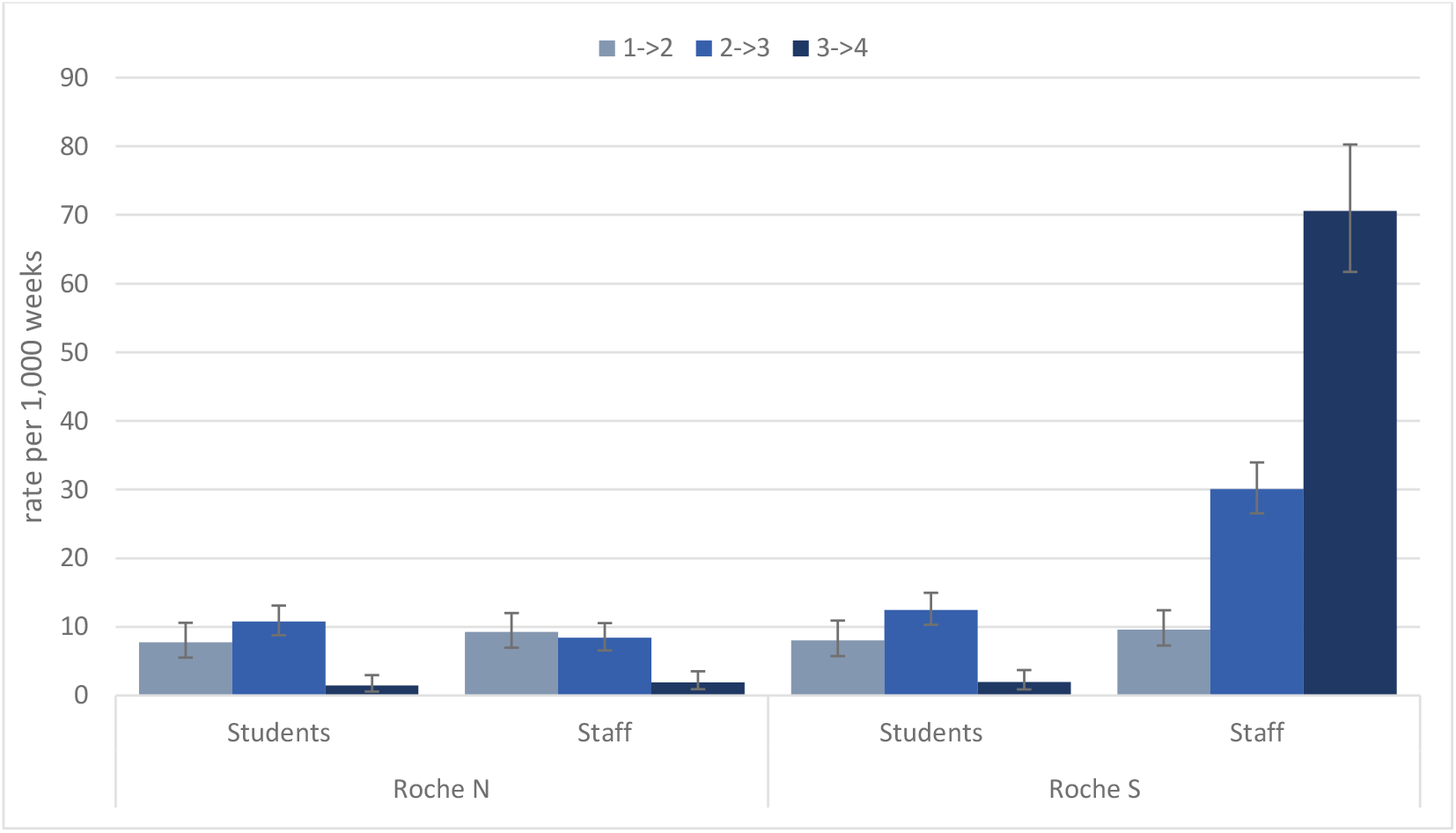
Seroconversion rates between testing rounds in students and staff using Roche N and S antibody results in student and staff sKIDs PLUS participants.

## Discussion

The full reopening of in-person schooling since March 2021 was associated with very few SARS-CoV-2 infections in urban secondary schools when assessed by both swabbing for active infection and blood sampling for SARS-CoV-2 antibodies, which captures both symptomatic and asymptomatic infection 2-4 weeks after infection. Reassuringly, by mid-July 2021, 97.8% of staff had SARS-CoV-2 antibodies, consistent with the successful national COVID-19 immunisation programme for adults. This compares with ∼40% of students who had acquired immunity through prior infection. Linkage with national RT-PCR testing data identified 12% of participants had confirmed infection during the 2020/21 academic year, with most infections occurring during the last 4 months of 2020, and infection rates closely following community infection rates at the time.

The UK situation provides unique insight into the impact of schools on community SARS-CoV-2 infection rates because schools remained open on at least two occasions when the rest of the country was in national lockdown. We previously reported that infection rates in school-aged children followed, but with a lag, adults in the community and declined following national lockdown in November 2020 despite schools remaining fully open at the time.^4^ The reopening of all schools on 08 March 2021 was associated with a small, transient increase in cases among school-aged children, driven mainly by whole-school LFD testing at the start of the term and twice-weekly home LFD testing for secondary school students. Cases did not increase in primary or secondary school students after they returned from their Easter holidays on 18 April until 17 May 2021, which coincided with the Step 3 easing of national lockdown. Despite the ready availability of testing for students, staff and their families, the proportion of undiagnosed infections, ascertained by comparing PCR diagnoses and antibody seroconversion rates, was high.

After 17 May 2021, the trajectory of cases in secondary-school aged students was similar to that of older teenagers and young adults who did not attend school, suggesting school re-opening was not a major factor driving the observed increase in community incidence. Rather, the easing of national lockdown likely provided more opportunities for mixing, and thereby transmission, among teenagers and young adults outside the school premises, which allowed for a more rapid spread of the virus. This would, consequently, lead to more introductions of the virus into educational settings, leading to mass educational disruption due to large numbers of students in bubbles self-isolating because of possible contact with a confirmed or suspected case.^20^

It should be noted that face coverings and masks were mandated in classrooms for secondary school students from 08 March 2021 until 17 May 2021 because of concerns about increased transmissibility of the Alpha variant, which was the predominant circulating variant at the time. Otherwise, face coverings in secondary schools in England were only recommended for staff and students in communal areas outside the classroom if physical distancing was difficult to maintain.^21^ The contribution of face masks in preventing infections in educational settings is uncertain, especially when compared to other school mitigations. One study found that the number of mitigations (face masks being one of them) was more important in reducing school infection rather than the individual mitigations *per se*.^22^ In some countries, children as young as 2 years are required to wear face coverings/masks in educational and care settings.^23-25^ There is a desperate need for robust studies assessing the benefits and harms of face coverings/masks in young children.^26^

The large increase in childhood COVID-19 cases since June 2021, with a similar trajectory as the Alpha variant in December 2021 followed by a high, but stable, plateau since mid-July 2021 suggests that SARS-CoV-2 antibody seroprevalence is likely to have increased significantly in secondary school aged students since our last sampling. In the UK, adults aged ≥18 years were offered COVID-19 vaccination from mid-June 2021, and the first dose of vaccine was offered to 16-17 year-olds from 04 August 2021.^27-29^ Vaccination of 12-15 year-olds in the UK, initially with a single dose only, started on 20 September 2021.^30^ Whilst acknowledging the limited additional direct benefits of vaccination in healthy teenagers who have a very low risk of severe COVID-19, it is hoped that vaccination will help reduce further educational disruption for students.^31^ Currently, schools are fully open in England and most mitigations including bubbles removed, with only unwell children and those with confirmed COVID-19 having to self-isolate from school.

### Strengths and limitations

The strength of this study is the longitudinal assessment of SARS-CoV-2 infection and transmission in secondary schools. One limitation was the large numbers of students self-isolating during the final sampling visit in June/July 2021. This will have contributed to the apparent low infection rates on Round 4 school testing day, but linkage with national testing data found only 1.5% of students who did not take part in Round 4 had a confirmed COVID-19, indicating that most students were self-isolating because they had been a contact of a case. Reassuringly, though, the lower number of participants tested in Round 4 will not have impacted on seroprevalence and seroconversion rates between Rounds 3-4 because of the delay in developing an antibody response after infection and, therefore, the antibody tests performed in Round 4 would reflect infection rates at least 2-4 weeks prior to testing. Given the continuing high SARS-CoV-2 infection rates in teenagers throughout the summer months and beyond, and with COVID-19 vaccination now recommended for teenagers, it is likely that our antibody positivity rates significantly underestimate current seroprevalence. Another limitation is that participation among staff also fell during the final round, mainly because most of them were vaccinated and no longer saw study participation as an opportunity to confirm their immunity status.

## Conclusions

The reopening of secondary schools since March 2021, whilst the rest of the country remained in national lockdown, was associated with very low rates of SARS-CoV-2 infection and antibody seroconversion until July 2021, despite the emergence and rapid spread of the Delta variant in England. Infection rates increased rapidly after the easing of national lockdown from 17 May 2021, however, this increase was too early to be detected in the final round of antibody sampling in July 2021. On-going serosurveillance in educational settings will be important to assess the impact of COVID-19 vaccination on infection and transmission in educational settings.

## Data Availability

Applications for relevant anonymised data should be submitted to the UK Health Security Agency Office for Data Release.

## Contributors

SNL, FB, JB, IOO, SA, JG, AJB, BB, GA, VS, KEB, and MER were responsible for conceptualisation and study design and methodology. SNL, FB, JB, IOO, SA, JG, AJB, BB, GI, FA, ZA-C, RB, EL, MZ, AL, JE, LW and JP contributed to project administration (including laboratory colleagues). SNL and GI contributed to the original draft of the report, did the formal analysis and were responsible for data validation and verification. All authors contributed to reviewing and editing of the manuscripts. All authors had access to the data; SNL and GI had final responsibility to submit for publication.

## Acknowledgments

We thank all those who contributed to the study; the PHE team, the schools, head teachers, staff, families, and their very brave children who took part in sKIDs PLUS.

## References

1. Viner RM, Mytton OT, Bonell C, et al. Susceptibility to SARS-CoV-2 Infection Among Children and Adolescents Compared With Adults: A Systematic Review and Meta-analysis. JAMA Pediatr 2021; 175(2): 143–56.

2. Ismail SA, Saliba V, Lopez Bernal J, Ramsay ME, Ladhani SN. SARS-CoV-2 infection and transmission in educational settings: a prospective, cross-sectional analysis of infection clusters and outbreaks in England. Lancet Infect Dis 2021; 21(3): 344–53.

3. Ladhani SN, Ireland G, Baawuah F, et al. SARS-CoV-2 infection, antibody positivity and seroconversion rates in staff and students following full reopening of secondary schools in England: A prospective cohort study, September–December 2020. EClinicalMedicine 2021; (In Press).

4. Mensah AA, Sinnathamby M, Zaidi A, et al. SARS-CoV-2 infections in children following the full re-opening of schools and the impact of national lockdown: Prospective, national observational cohort surveillance, July-December 2020, England. J Infect 2021; 82(4): 67–74.

5. Public Health England. Weekly national Influenza and COVID-19 surveillance report-Week 26 report (up to week 25 data), 2021.

6. Public Health England. SARS-CoV-2 variants of concern and variants under investigation in England-Technical briefing 17, 2021.

7. Lopez Bernal J, Andrews N, Gower C, et al. Effectiveness of Covid-19 Vaccines against the B.1.617.2 (Delta) Variant. N Engl J Med 2021; 385(7): 585–94.

8. Office for National Statistics. Coronavirus (COVID-19) Infection Survey, UK: 24 September 2021, 2021.

9. Department of Health and Social Care. Policy paper: UK COVID-19 vaccines delivery plan, 2021.

10. Department for Education. Mass testing for secondary pupils as all schools and colleges fully reopen from 8 March. 2021.

11. Ladhani SN, Ireland G, Baawuah F, et al. Emergence of SARS-CoV-2 Alpha (B.1.1.7) variant, infection rates, antibody seroconversion and seroprevalence rates in secondary school students and staff: Active prospective surveillance, December 2020 to March 2021, England. J Infect 2021.

12. Ladhani SN, Amin-Chowdhury Z, Amirthalingam G, Demirjian A, Ramsay ME. Prioritising paediatric surveillance during the COVID-19 pandemic. Arch Dis Child 2020; 105(7): 613–5.

13. Tasso. Tasso+. 2021. https://www.tassoinc.com/tasso-plus (accessed 28 September 2021 2021).

14. Niu P, Liu R, Zhao L, et al. Three Novel Real-Time RT-PCR Assays for Detection of COVID-19 Virus. China CDC Wkly 2020; 2(25): 453–7.

15. Corman VM, Landt O, Kaiser M, et al. Detection of 2019 novel coronavirus (2019-nCoV) by real-time RT-PCR. Euro Surveill 2020; 25(3).

16. Public Health England (PHE). Evaluation of the Abbott SARS-CoV-2 IgG for the detection of anti-SARS-CoV-2 antibodies. 08 June 2020. https://assets.publishing.service.gov.uk/government/uploads/system/uploads/attachment_data/file/890566/Evaluation_of_Abbott_SARS_CoV_2_IgG_PHE.pdf.

17. Public Health England. Evaluation of the Abbott SARS-CoV-2 IgG for the detection of anti-SARSCoV-2 antibodies, 2020.

18. Harris RJ, Whitaker HJ, Andrews NJ, et al. Serological surveillance of SARS-CoV-2: Six-month trends and antibody response in a cohort of public health workers. J Infect 2021.

19. NHS Digital. The Personal Demographics Service. England; 2021.

20. Weale S. Covid-related pupil absences in England jump to 840,000. The Guardian. 2021 13/07/2021.

21. Department for Education. Guidance for full opening: schools. 2020.

22. Lessler J, Grabowski MK, Grantz KH, et al. Household COVID-19 risk and in-person schooling. Science 2021; 372(6546): 1092–7.

23. Sandy Ong. Should young children be made to wear face masks? BBC. 2021 25/10/2021.

24. Alec Fenn. Should children wear face masks at primary school? CGTN. 2021 02/11/2021.

25. Todd J. Why Having Your Toddler Wear A Face Mask Could Make A Big Difference. CBS Denver. 2021 17/08/2021.

26. Eberhart M, Orthaber S, Kerbl R. The impact of face masks on children-A mini review. Acta Paediatr 2021; 110(6): 1778–83.

27. Department of Health and Social Care. Independent report: JCVI statement on COVID-19 vaccination of children and young people aged 12 to 17 years: 4 August 2021, 2021.

28. NHS England. NHS invites all adults to get a COVID jab in final push. 2021.

29. NHS England. 21 and 22 year olds to be offered COVID-19 jab from today. 2021.

30. NHS England. One million children and young people can get NHS COVID jab. 2021.

31. Department of Health & Social Care. Research and analysis: Impact on school absence from COVID-19 vaccination of healthy 12 to 15 year old children, 2021.

