## Supplementary Materials for "Emergence of the Delta Variant and risk of SARS-CoV-2 infection in secondary school students and staff: prospective surveillance in 18 schools, England"

### *Supplementary Table 1: Predictors of the number of rounds students and staff participated in*

| | Students | | | Staff | | |
|---|---|---|---|---|---|---|
| | Odds ratio | 95% CI | p-value | Odds ratio | 95% CI | p-value |
| **Sex** | | | | | | |
| Female | 1 | | | 1 | | |
| Male | 1.01 | 0.80-1.26 | 0.96 | 1.15 | 0.89-1.49 | 0.28 |
| **Age cat** | | | | | | |
| Lower school | 1 | | | | | |
| GCSEs | 0.66 | 0.51-0.84 | 0.0015 | | | |
| A-Levels/College | 0.72 | 0.53-0.97 | | | | |
| 19-29 | | | | 1.4 | 0.99-1.98 | |
| 30-39 | | | | 1 | | 0.0022 |
| 40-49 | | | | 1.66 | 1.20-2.28 | |
| 50-59 | | | | 1.76 | 1.28-2.43 | |
| 60+ | | | | 2.07 | 1.16-3.68 | |
| **Ethnicity** | | | | | | |
| White | 1 | | | 1 | | |
| Black | 0.98 | 0.60-1.62 | | 0.73 | 0.42-1.27 | |
| Asian | 0.72 | 0.54-0.96 | 0.19 | 0.75 | 0.52-1.09 | 0.46 |
| Mixed | 0.80 | 0.54-1.19 | | 1.20 | 0.59-2.46 | |
| Other | 1.10 | 0.64-1.90 | | 1.03 | 0.40-2.66 | |
| **School area** | | | | | | |
| Derbyshire | 1 | | | 1 | | |
| East London | 1.42 | 1.04-1.93 | | 0.48 | 0.35-0.67 | |
| Greater Manchester | 1.05 | 0.71-1.57 | | 0.57 | 0.35-0.94 | |
| Hertfordshire | 0.91 | 0.59-1.40 | <0.0001 | 1.78 | 1.03-3.06 | <0.0001 |
| West London | 1.96 | 1.39-2.77 | | 0.90 | 0.60-1.33 | |
| Birmingham | 2.61 | 1.84-3.69 | | 1.68 | 1.20-2.35 | |

***Supplementary Figure 2: SARS-CoV-2 positivity, and 95% confidence intervals, in student and staff participants over 4 rounds of secondary school testing.***

*In round 3 no staff tested positive for SARS-CoV-2.*

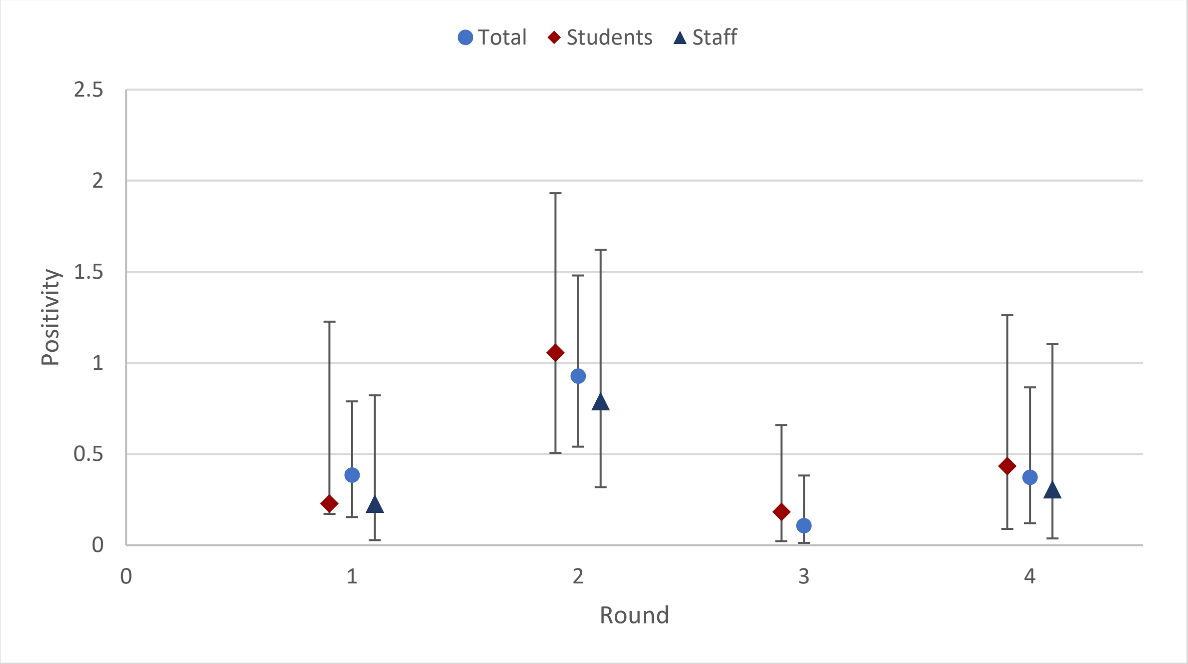

***Supplementary Figure 3: a) Week of SARS-CoV-2 infection, from study testing and SGSS reported diagnoses, in student and staff sKIDs PLUS participants over the study period; b) COVID-19 Infection Survey 14-day weighted estimates of the percentage of 11/12 to 15/16 year olds testing positive for COVID-19 in England; c) diagnosed weekly COVID-19 case rates per 100,000 population by age; d) variant type as a percent of all samples genotyped in England between 1 Feb and 7 August 2021***

a)

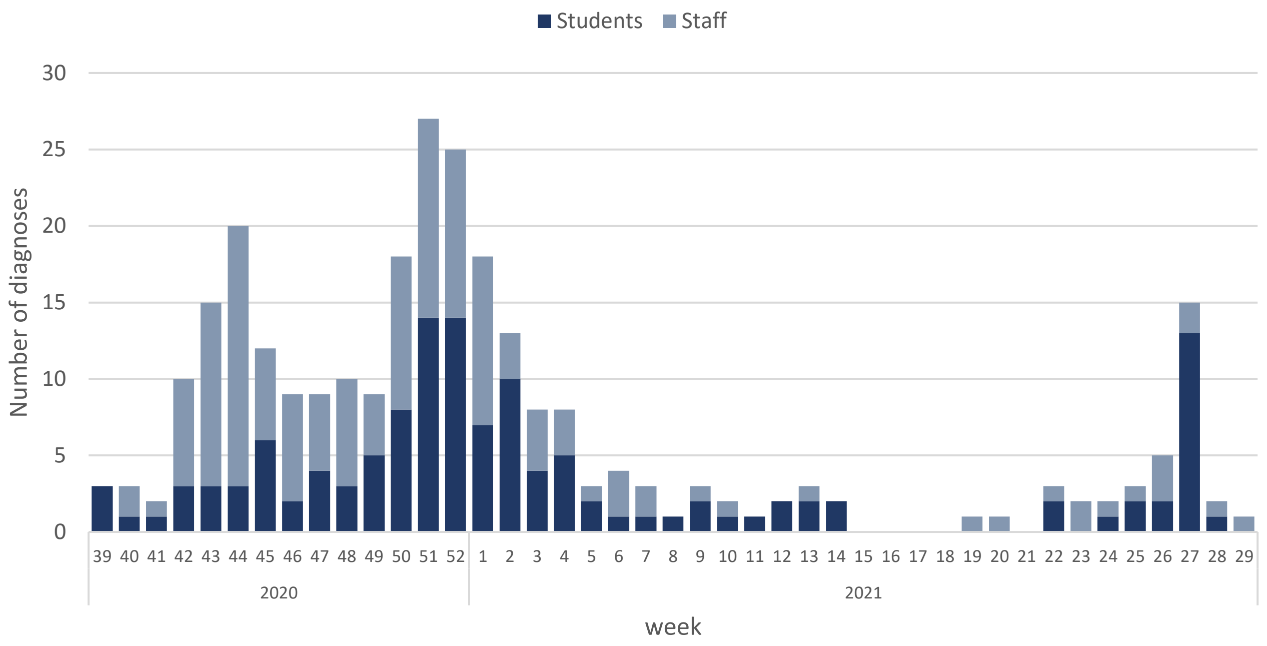


b)

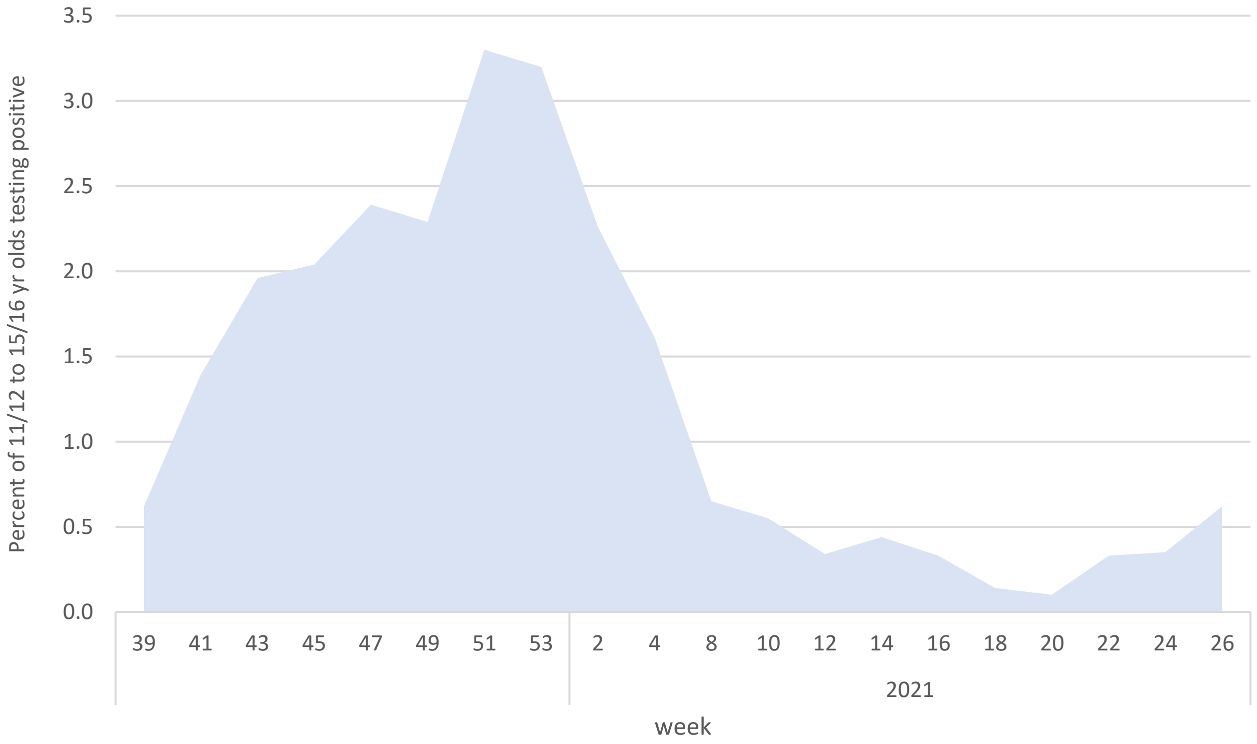

c)

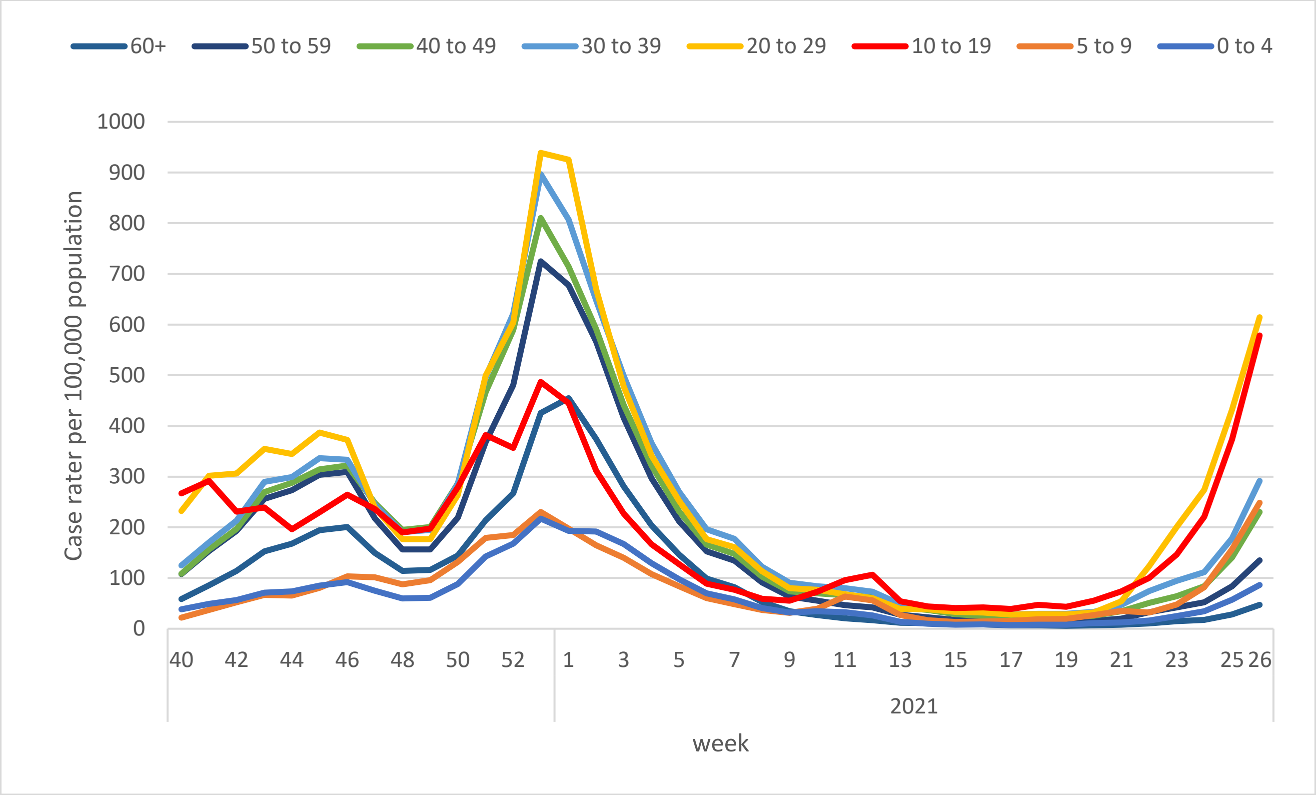
60+
50 to 59
40 to 49
30 to 39
20 to 29
10 to 19
5 to 9
0 to 4
Case rater per 100,000 population
1000
900
800
700
600
500
400
300
200
100
0
40 42 44 46 48 50 52 1 3 5 7 9 11 13 15 17 19 21 23 25 26
2021
week

d)

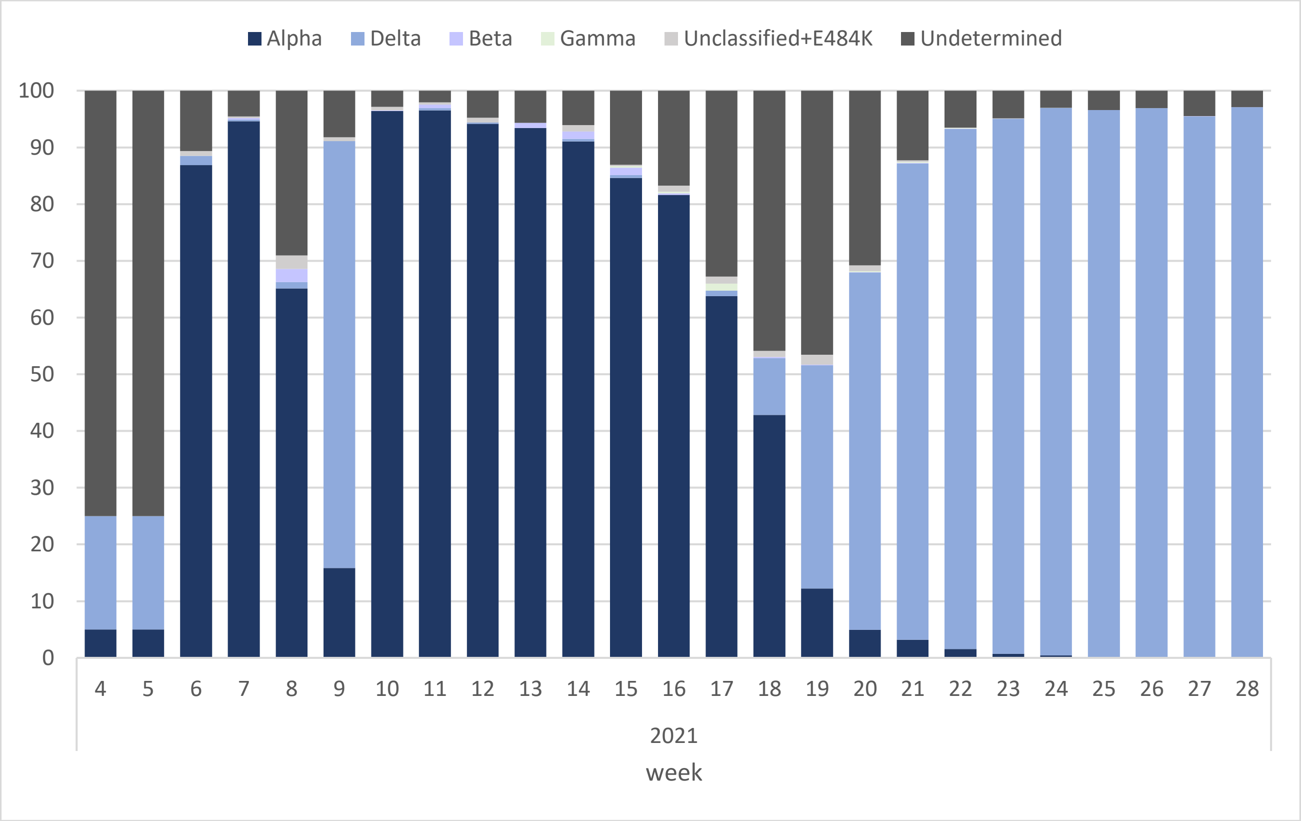
Alpha
Delta
Beta
Gamma
Unclassified+E484K
Undetermined
100
90
80
70
60
50
40
30
20
10
0
4 5 6 7 8 9 10 11 12 13 14 15 16 17 18 19 20 21 22 23 24 25 26 27 28
2021
week

***Supplementary Figure 4: Roche N and S seroprevalence in sKIDs Plus students and staff by school area and compared to 4-week community (regional) estimates from NHSBT blood donor seroprevalence surveillance in round 1 (a), round 2 (b), round 3 (c) and round 4 (d).***

*Students are compared to 18-30 year old blood donors and staff to 18-64 year old donors. 4-week dates were; 21 Sept-18 Oct 2020 for round 1, 23 Nov-20 Dec 2020 for round 2, 15 Mar-11 Apr 2021 for round 3 and 28 Jun-25 Jul 2021 for round 4.Community S estimates are not available for round 1 and were not used in round 4 for students due to high vaccination levels in this population.*

a)

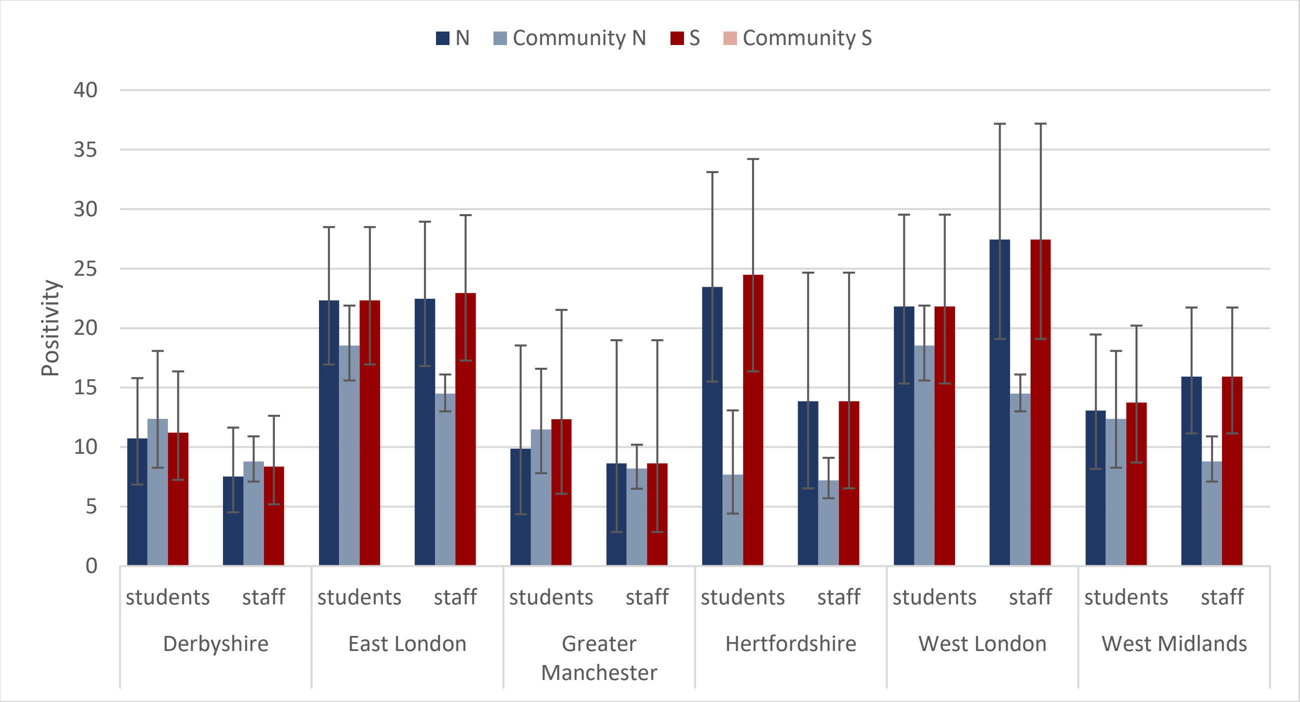


b)

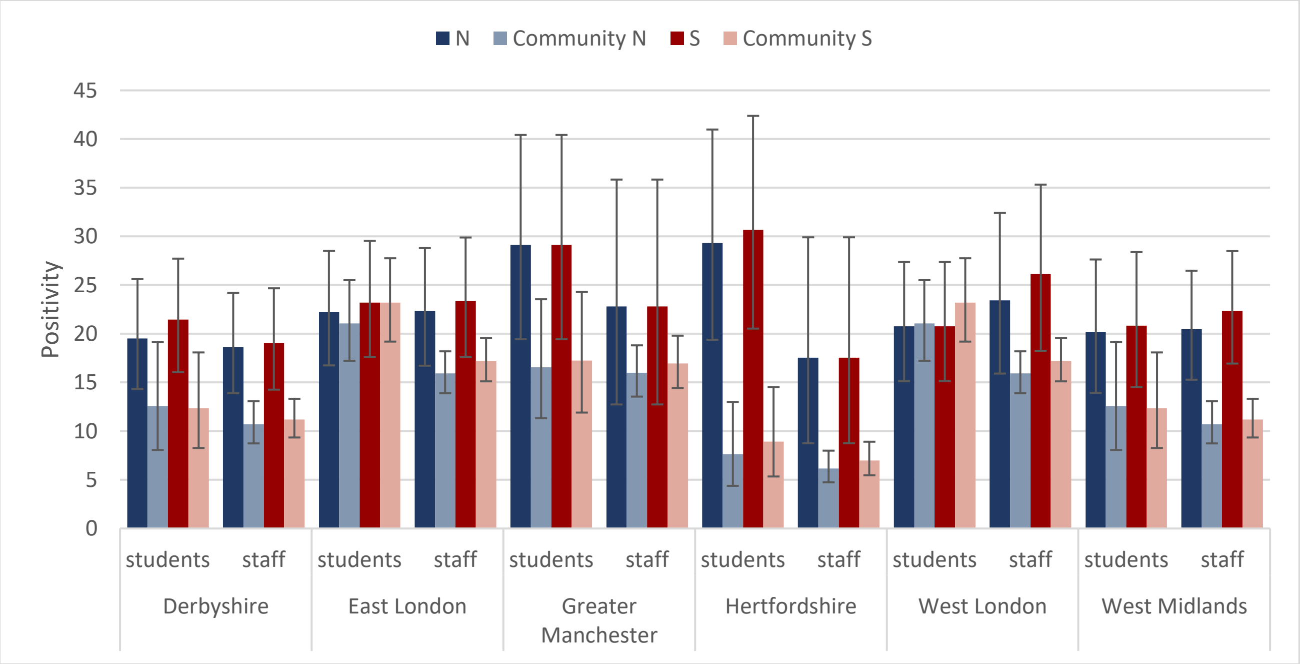

c)

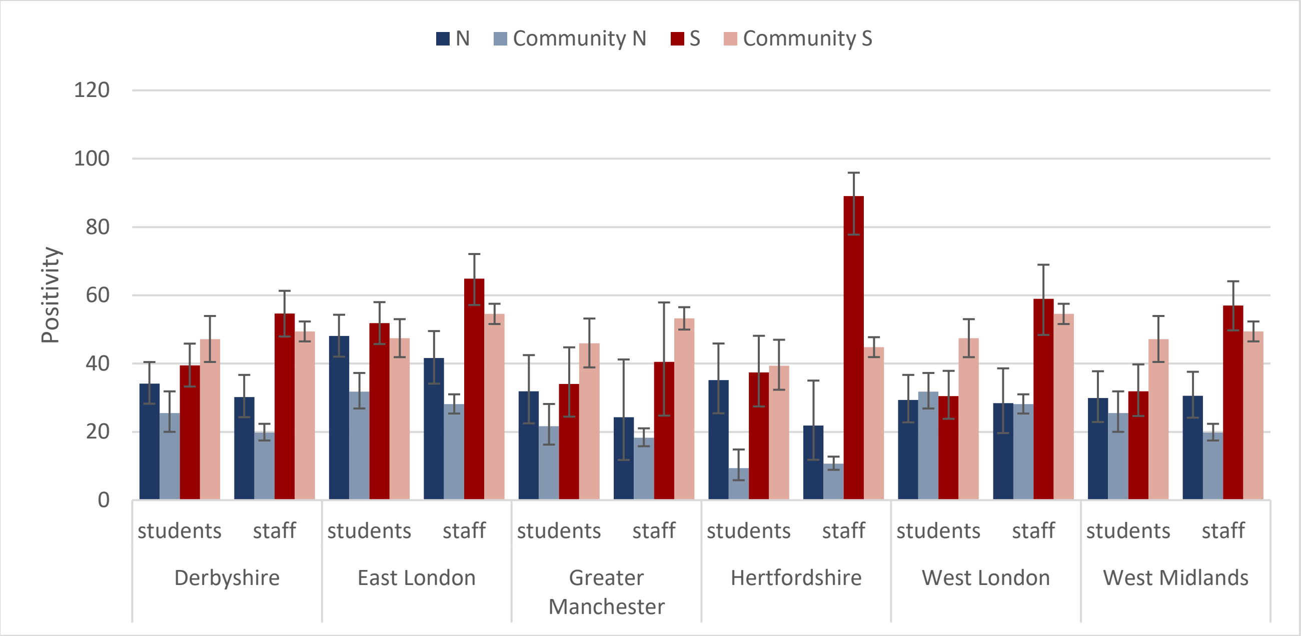
N
Community N
S
Community S
Positivity
120
100
80
60
40
20
0
students
staff
Derbyshire
East London
Greater Manchester
Hertfordshire
West London
West Midlands

d)

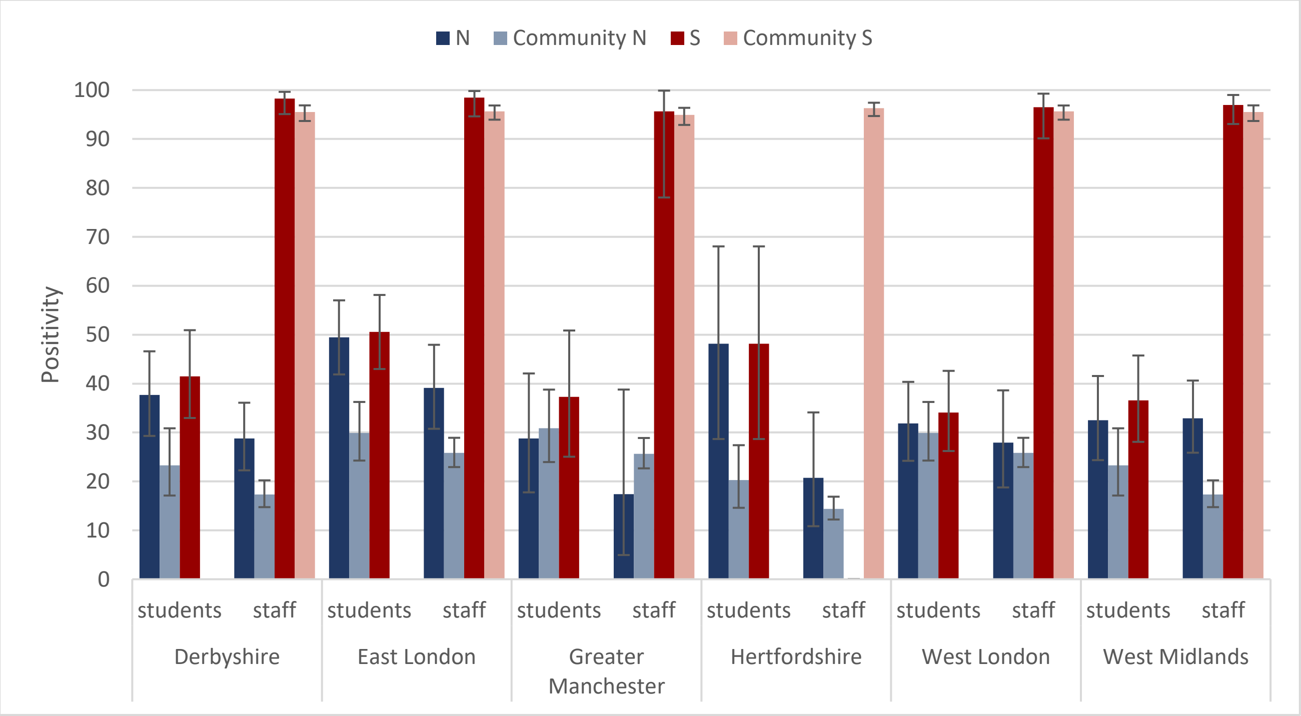
N
Community N
S
Community S
Positivity
100
90
80
70
60
50
40
30
20
10
0
students
staff
Derbyshire
East London
Greater Manchester
Hertfordshire
West London
West Midlands

***Supplementary Table 5: N and S antibody seroconversion in sKIDs Plus participants who were sampled in consecutive rounds on the Roche N assay (a) and Roche S assay (b).***

a)

| | | Sample | Seroconverters | | | |
|---|---|---|---|---|---|---|
| | | | N | Percent | Rate per 1,000 weeks | 95% CI |
| **Round 1->2** | | | | | | |
| | Total | 1131 | 94 | 8.3 | 8.56 | 6.92-10.46 |
| | Students | 523 | 39 | 9.0 | 7.76 | 5.52-10.59 |
| | Staff | 608 | 55 | 7.5 | 9.24 | 6.97-12.01 |
| **Round 2->3** | | | | | | |
| | Total | 1128 | 172 | 15.2 | 9.63 | 8.25-11.17 |
| | Students | 584 | 100 | 17.1 | 10.78 | 8.78-13.10 |
| | Staff | 544 | 72 | 13.2 | 8.39 | 6.57-10.55 |
| **Round 3->4** | | | | | | |
| | Total | 734 | 17 | 2.3 | 1.69 | 0.98-2.70 |
| | Students | 350 | 7 | 2.0 | 1.44 | 0.58-2.97 |
| | Staff | 384 | 10 | 2.6 | 1.92 | 0.92-3.53 |

b)

| | | Sample | Seroconverters | | | |
|---|---|---|---|---|---|---|
| | | | N | Percent | Rate per 1,000 weeks | 95% CI |
| **Round 1->2** | | | | | | |
| | Total | 1124 | 97 | 8.6 | 8.89 | 7.21-10.83 |
| | Students | 518 | 40 | 7.7 | 8.03 | 5.74-10.92 |
| | Staff | 606 | 57 | 9.4 | 9.60 | 7.28-12.42 |
| **Round 2->3** | | | | | | |
| | Total | 1111 | 368 | 33.1 | 20.93 | 18.86-23.15 |
| | Students | 576 | 114 | 19.8 | 12.47 | 10.30-14.96 |
| | Staff | 535 | 254 | 47.5 | 30.08 | 26.54-33.95 |
| **Round 3->4** | | | | | | |
| | Total | 553 | 223 | 40.3 | 29.19 | 25.53-33.22 |
| | Students | 331 | 9 | 2.7 | 1.95 | 0.89-3.71 |
| | Staff | 222 | 214 | 96.4 | 70.56 | 61.70-80.26 |
